# Heterologous vaccination strategy for containing COVID-19 pandemic

**DOI:** 10.1101/2021.05.17.21257134

**Authors:** Ang Lin, JingJing Liu, Xiaopin Ma, Fanfan Zhao, Bo Yu, Jiaxin He, Mingyun Shen, Lei Huang, Hongming Tang, Erpeng Jiang, Yue Wang, Pingfang Cui, Yujian Zhang, Weiguo Yao, Aihua Zhang, Youchun Wang, Yuhua Li, Weijin Huang, Qihan Li, Zhongmin Liu, Hangwen Li

## Abstract

An unequitable vaccine allocation and continuously emerging SARS-CoV-2 variants pose challenges to contain the pandemic, which underscores the need for licensing more vaccine candidates, increasing manufacturing capacity and implementing better immunization strategy. Here, we report data from a proof-of-concept investigation in two healthy individuals who received two doses of inactivated whole-virus COVID-19 vaccines, followed by a single heterologous boost vaccination after 7 months with an mRNA vaccine candidate (LPP-Spike-mRNA) developed by Stemirna Therapeutics. Following the boost, Spike-specific antibody (Ab), memory B cell and T cell responses were significantly increased. These findings indicate that a heterologous immunization strategy combining inactivated and mRNA vaccines can generate robust vaccine responses and therefore provide a rational and effective vaccination regimen.

## Introduction

Inoculation of COVID-19 vaccines has been rolling out globally, but current vaccine supply cannot suffice unprecedented global need and has caused unfair vaccine access in low/middle-income nations^1^. While collaborative efforts have been made to ensure equitable allocation, it is far less likely to reach this goal as opposed to the high contagion speed. Just in the time of finalizing this study, The World Health Organization (WHO) approved the emergency use of an inactivated vaccine (BBIBP-CorV) developed by Sinopharm Group Co., Ltd.^2^, which could potentially help relieve the global vaccine shortfall pressure.

Although a few COVID-19 vaccines developed from different technologies have been approved based on the efficacy data from current clinical trials, the duration of protection is still unclear and remains to be continuously monitored. Emergence of SARS-CoV-2 variants with increased transmissibility or infectivity are becoming endemically dominant and showed evidence to impair vaccine efficacy at a varing level^3-5^. This urges the development of next-generation variant-matched vaccines, multivalent vaccines or optimization of current vaccination strategy, for example, by introducing a booster dose following primary immunization series. In most cases, multiple doses of vaccines are required to elicit long-lasting and efficient protection. Homologous prime/boost vaccination strategy has widely been applied and shown potency to enhance humoral responses^6^. Similar to this regimen, heterologous prime/boost vaccination strategy that combines different vaccine platforms has gained significant momentum during past decades and proven to be more effective at improving vaccine efficacy in pre-clinical studies, but yet been licensed for human^7^.

In this report, we evaluated the vaccine responses in two healthy individuals that were fully vaccinated with inactivated COVID-19 vaccines and 7 months later boosted with a novel COVID-19 mRNA vaccine (LPP-Spike-mRNA).Following the booster, both two subjects demonstrated excellent safety profile with no fever or any other obvious systemic and/or local adverse symptoms, as well as normal blood biochemistry parameters. In addition, they showed rapid and strong vaccine responses with regard to a significant increase in antigen-specific binding IgG, neutralizing Abs (NAbs), Spike-specific IgG^+^ memory B cells as well as Spike-specific IFN-γ, IL-2, or IL-21-producing T cells. This indicates that heterologous vaccination, in this case an inactivated whole-virus vaccine for two priming doses and an mRNA vaccine for booster, may be a strategy in the ongoing mass vaccination and would add to the prospect of controlling the pandemic.

## Methods

### Study subjects, immunization and sample collection

This study was approved by the local ethical committee at Tongji University East Hospital, Shanghai, China, and was performed according to the Declaration of Helsinki Principles. Two subjects were enrolled in this study and gave written informed consent. Subject 1 was a man and subject 2 was a woman. Both two subjects are at the age between 50-55. They participated in a Phase I clinical trial on an inactivated whole-virus COVID-19 vaccine (ClinicalTrial.gov: NCT04412538), where they received two doses (100EU) of vaccines at an interval of 28 days. This inactivated vaccine was developed by the Institute of Medical Biology (IMB), Chinese Academy of Medical Sciences (CAMS) and is currently under evaluation in a Phase III trial (ClinicalTrial.gov: NCT04659239). Around 7 months after the second immunization, they were administered with a novel COVID-19 mRNA vaccine (25 mcg) developed by Stemirna Therapeutics, which is currently under clinical evaluation in China. Prior to and following the boost vaccination, peripheral venous blood was collected in EDTA vacuum tubes or Vacutainer Serum tubes (BD) for the prepration of perihperal blood mononuclear cells (PBMCs) and sera samples, respectively. PBMCs were isolated using Ficoll-Pague PLUS density gradient solution (GE Healthcare) as previously described^8^. For comparison of Ab response, sera collected from 15 convalescent COVID-19 patients were assessed using same method. These patients had a history of PCR-confirmed SARS-CoV-2 infection 1 to 3 months before sample collection.

### Binding IgG measurement

Titers of binding IgG specific for pre-fusion structured Spike protein, Spike protein ectodomain (S-ECD) and receptor binding domain (RBD) were evaluated by enzyme-linked immunosorbent assay (ELISA). Antigens (Genscript) diluted in coating buffer (Biolegend) were coated into 96- well plates (Greiner Bio-One) at a concentration of 100ng/well and incubated overnight at 4□. The plates were then washed with PBS containing 0.05% Tween-20 (PBST) and subsequently blocked with 2% bovine serum albumin (BSA) for 2 h at 25□. Following this, serum samples serially diluted in PBST containing 0.2% BSA were added and incubated for 2h at 25□. Binding IgG was evaluated using HRP-conjugated sheep anti-human IgG Ab (1:50,000) for 1 hour at 25□. TMB Substrate was used for development, and the absorbance was read at 450 nm (minus 610 nm for wavelength correction). Endpoint titer was calculated as the dilution that emitted an optical density (OD) value above 2.1× background. When background OD value is less than 0.05, take 0.05 for calculation.

### Neutralizing Ab measurement

Levels of neutralizing Abs (NAbs) were measured using three different approaches including pseudotyped virus neutralization test (pVNT), surrogate virus neutralization test (sVNT) and cytopathic effect (CPE) assay using live SARS-CoV-2 virus. pVNT assay was performed as previously reported^9^ where in-house constructed vesicular stomatitis virus (VSV) expressing SARS-CoV-2 Spike protein (strain Wuhan-Hu-1) is used to infect ACE2-expressing Huh7 cells. CPE assay was performed in Biosafety Level 3 (BSL-3) laboratory at the Chinese Academy of Medical Sciences according to a previously described protocol^10^. sVNT assay was developed according to a reported protocol with modifications^11^, which is based on the rationale that NAbs can block the interaction between RBD and ACE2. In brief, HRP-conjugated RBD (500ng/ml) and serially diluted serum were pre-incubated for 30 minutes at 37□. HRP-RBD incubated with 0.2% BSA was used as negative control. A 100-μL mixture was then transfered to 96-well plates pre-coated with hACE2 (100ng/well) for a 15-minute incubation at 37□. Following intensive washing, unbound HRP-RBD antigens were removed. TMB Substrate was used for development, and the absorbance was read at 450 nm (minus 610 nm for wavelength correction). For detemination of ID_50_ titer, RBD-ACE2 binding inhibition rate (%) was first calculated following the formula: (1- sample OD value/negative control OD value) × 100, followed by determination using Reed-Muench method.

### Enzyme linked immunospot (ELISpot) assay

Fequencies of different types of antigen-specific T cells were quantified by ELISpot assay, using Human IFN-γ, IL-2 or IL-21 ELISpot^plus^ Kits (Mabtech, Sweden) according to the manuals. For cell stimulation, S-ECD protein (10μg/ml) or RBD (10μg/ml) were used to stimulate 3×10^5^ PBMCs for 20 hours prior to the following detection. Spots were developed with BCIP/NBT substrate solution and counted using Immunospot S6 analyzer (CTL).

### Memory B cell analysis

Frequency of Spike-specific memory B cells was assessed by flow cytometry. For the preparation of probes, biotinylated Spike proteins were conjugated with PE- or APC-labelled streptavidin at a molar ratio of 4:1. One million PBMCs were first incubated with probes for 20 minutes at 4□, and then stained with LIVE/DEAD Fixable Aqua Dead Cell Stain Kit (BD) and antibody cocktails for 20 minutes at 4□ in the dark. Flow cytometric analysis was carried out on FACS Canto™ II cell analyzer (BD Biosciences). Data were analyzed using FlowJo V.10.1 (Tree Star). Antibody cocktail contains the following fluorescently-labelled antibodies: anti-human CD3 Ab (clone: SP34-2), anti-human CD8 Ab (clone: RPA-T8), anti-human CD14 Ab (clone: M5E2), anti-human CD16 Ab (clone: 3G8), anti-human CD20 Ab (clone: 2), anti-human IgM Ab (clone: G20-127), anti-human IgG Ab (clone: G18-145).

## Results

The study design is illustrated in Figure 1A. The magnitude and kinetics of Ab responses in the two subjects were first evaluated. The day of the boost vaccination (day 0), low but detectable levels of Spike-specific IgG were present as the result of durability of the responses generated by the prime vaccinations with the inactivated vaccine as expected. After the mRNA vaccine booster, the levels of IgG directed to pre-fusion Spike, S-ECD as well as RBD showed a rapid and robust increase (Figure 1B). The levels peaked at day 14 reaching reciprocal titers of 51200 in both subjects, which far exceeded the geometric mean titers (GMTs) of binding IgG in convalescent serum from individuals who had COIVD-19, which are 7699 (to pre-fusion S), 8844 (to S-ECD) and 4031 (to RBD), respectively. The neutralizing capacity of the vaccine-induced Abs was next assessed by three complementary assays using either pseudovirus, live virus or virus-free system. In line with the strong induction of Spike-binding IgG, the levels of NAbs were also significantly augmented upon the mRNA vaccine booster, reaching 2457, 111 and 256 reciprocal ID_50_ GMT at day 14 as detected by pVNT, sVNT and CPE assay, respectively (Figure 1C-E). Again, the magnitude was markedly higher than that detected in convalescent samples regardless of assay.

**Figure 1.**
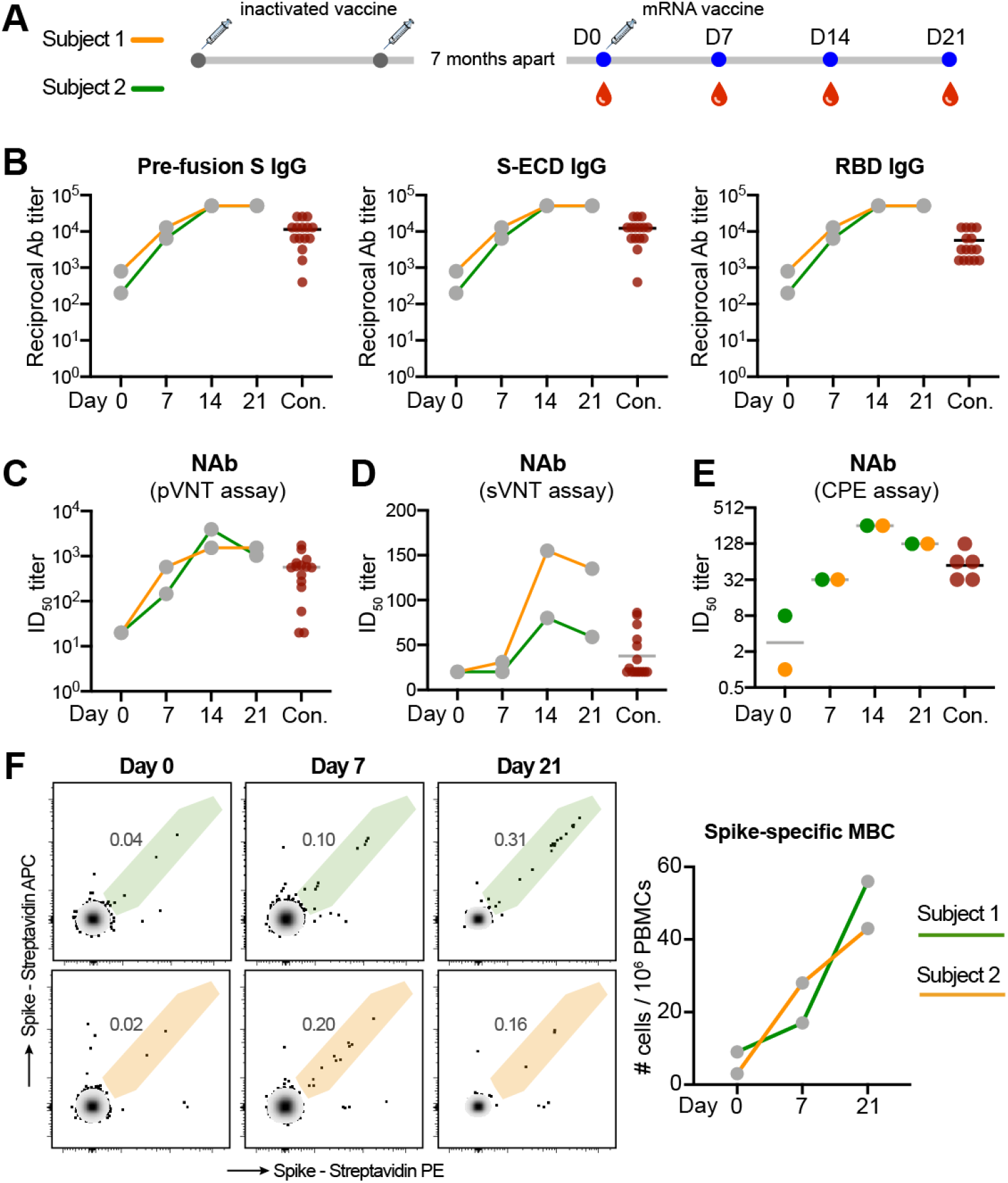
Antibody and memory B cell responses after a heterologous COVID-19 vaccination. Panel A. Two subjects received a heterologous prime/boost vaccination combining an inactivated vaccine and an mRNA vaccine as depicted. Panel B. Levels of antigen-specific binding IgG were assessed on day 0 and day 7, 14, 21 following the mRNA vaccine booster. Panel C-E. Neutralizing ability of Abs were measured using pVNT, sVNT, and CPE assay. The 50% inhibitory dilution (ID50) titer is shown. Panel F. Frequencies of Spike protein-specific IgG^+^ memory B cells are shown. Cells are gated on CD20^+^IgD^-^IgM^-^ class-switched B cells.

Successful vaccination should be able to elicit strong immunological memory that can react immediately upon a secondary antigen exposure. We therefore followed the induction of memory B cells after vaccination. 7 months after the second dose of the inactivated vaccine, a small population of Spike-specific IgG^+^ memory B cells were still detectable in the blood circulation of both subjects (Figure 1F), indicating the induction and maintenance of antigen-specific memory B cells. However, the Spike-specific memory B cell pool was further expanded upon the mRNA vaccine booster, as evidenced by a clear increase of specific memory B cells (Figure 1F, right panel).

The phenotype and magnitude of Spike-specific T cells were also analyzed. Prior to the booster, both the two subjects had low levels of IFN-γ-secreting T cells reacting with SARS-CoV-2 antigens (Figure 2A-B) as part of the response to the prime immunizations with the inactivated vaccine. The mRNA vaccine booster elicited a strong Th1-polarized T cell response as evidenced by the increase of IFN-γ or IL-2-secreting T cells upon S-ECD or RBD stimulation (Figure 2A-B). In contrast, no or very few IL-4-secreting T cells were detected (data not shown). In addition to these, a strong induction of IL-21-secreting T cells was observed upon antigen exposure. The presence of IL-21 is indicative of a T follicular helper (Tfh) cell response that could contribute to the generation of memory B cells and high-quality Abs^12^ found with the mRNA vaccine booster (Figure 1).

**Figure 2.**
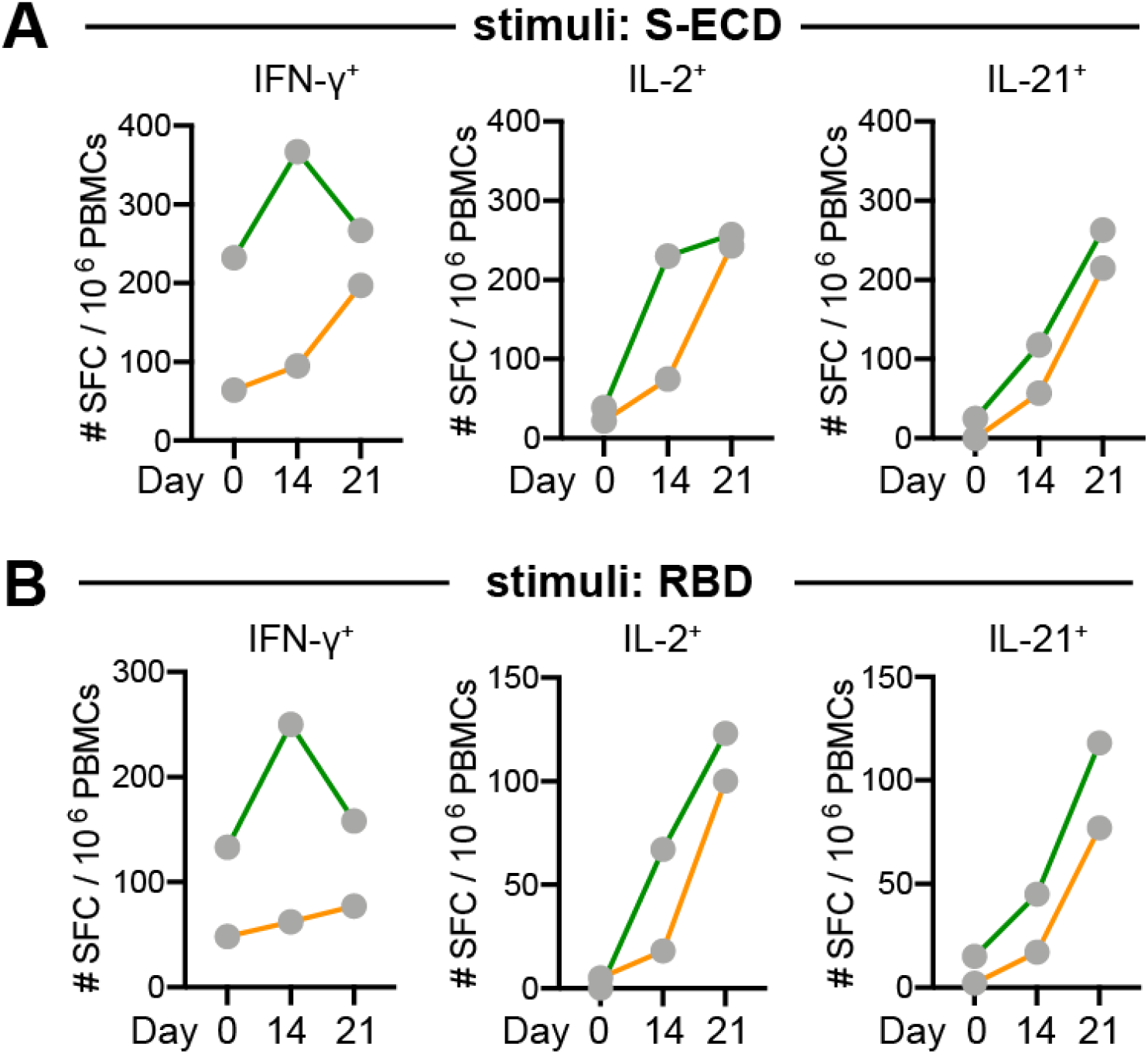
Antigen-specific T cell responses after a heterologous COVID-19 vaccination. PBMCs isolated at day 0 and day 14, 21 after the booster were stimulated with S-ECD (Panel A) or RBD (Panel B) at a concentration of 10 μg/ml for 20 hours. Frequencies of IFN-γ, IL-2 or IL- 21-secreting T cells were evaluated by ELISpot and are enumerated as spot-forming cells (SFC) per million stimulated PBMCs.

## Discussion

As of May 7, 2021, at least 14 vaccines have been approved for full or emergency use worldwide, with six of them being approved by WHO. However, it has become clear that in this high demand of vaccines that more vaccine candidates are likely needed as well as information on how combination of different vaccines can be given. Also, we are still lacking knowledges on some key questions regarding these novel vaccines. Different types of vaccines differ in their mechanisms of action and generate differential vaccine outcomes in terms of magnitude, phenotype, functionality, durability, and most importantly the protective efficacy. Specific characteristics of vaccines especially the ones developed from new platforms, such as mRNA vaccines, remain largely unknow as no such vaccines have been licensed for human use before the pandemic. Clinical evidence has indicated a potentially rapid waning immunity in COVID-19 convalescent patients or immunized individuals^13,14^, highlighting the necessity to recieve COVID-19 vaccination routinely similar to seasonal influenza vaccination. In addition, the emergence of SARS-CoV-2 variants showing vaccine resistance are dominating endemically and have raised concerns on the loss of efficacy from the currently-used vaccines developed based on early virus isolates^15^. These challenges have underscored the need for developing novel or improved vaccines, for example a universal variant-proof vaccine, or in a more timely and efficient way, implementing an optimal vaccination strategy to boost vaccine responses exceeding protective threshold for all variants.

Induction and maintenance of long-lasting protection can be achieved through repeated doses of same vaccines called homologous prime/boost vaccination strategy. Alternatively, the heterologous prime/boost strategy based on sequential vaccination with two different vaccine platforms represents a new way of immunization and has been investigated in pre-clinical or clinical studies, which proven to be more efficient at enhancing both Ab and cellular immunity^7^. However, very limited data are available regarding the efficacy of heterologous strategy for COVID-19 vaccines in humans. In this report, we provided original evidence from two subjects receiving heterologous vaccination with the inactivated COVID-19 vaccine followed by boost with a novel mRNA vaccine (LPP-Spike-mRNA). Spike-specific Abs, memory B cell and T cell responses in these two subjects were significantly augmented by the heterologous boost. The elicited binding IgG and NAb titers were much higher than that in convalescent samples from individuals recovered from COVID-19. In addition, T cell responses in the two subjects were significantly elevated and demonstrated a dominantly Th1-polarized phenotype. Since inactivated vaccines adjuvanted with aluminium are more prone to elicit a Th2-type immune response, using mRNA vaccine as a boost offers the possibility to skew the phenotype towards Th1 that is more favorable for protection against SARS-CoV-2^16^. In addition, IL-21-producing antigen-specific T cells were strongly induced, which indicated a robust induction or activation of Tfh cells upon boost, although the cell surface phenotype was not confirmed directly in this study.

In line with our observations, a recent study^17^ has comprehensively evaluated the outcomes of heterologous vaccination using different types of COVID-19 vaccines in mice and found that a significantly enhanced vaccine response was achieved by heterologous vaccination as compared with the homologous approach. Therefore, heterologous vaccination strategy holds the prospect for augmenting vaccine efficacy.The limitation in our study is the lack of control subjects receiving a homologous booster with the same inactivated vaccine for a side-by-side comparison. Therefore the findings we presented here remain to be further validated in large cohort of subjects. In addition, other parameters need to be investigated in the context of heterologous COVID-19 vaccination, for example different vaccine types, order of vaccine administration, interval between priming and booster, and number of doses. A very recently initiated clinical trial (ClinicalTrial.gov: NCT04833101) aiming to study the outcomes of heterologous COVID- 19 vaccination using a viral-vector based and a RBD-based subunit vaccine will offer a good opportunity for further investigations on this topic.

## Data Availability

The data that support the findings of this study are available on request from the corresponding author.

## Acknowledgement

We thank the two volunteers for participating this study and the staff at Shanghai East Hospital, Tongji University for assistance with clinical trial operation and sampling. The study was supported by the National Key Research and Development Program of China (2020YFC0860300, to H.L), Shanghai Pujiang Talent Program (2020PJD068, to A.L), Shanghai Scientific Development Project (20431900300, to H.L) and internal funds from Stemirna Therapeutics, Inc.

